# CEREBLEED: Automated quantification and severity scoring of intracranial hemorrhage on non-contrast CT

**DOI:** 10.1101/2025.06.13.25329546

**Authors:** Santiago Cepeda, Olga Estaban-Sinovas, Ignacio Arrese, Rosario Sarabia

## Abstract

**Background:** Intracranial hemorrhage (ICH), whether spontaneous or traumatic, is a neurological emergency with high morbidity and mortality. Accurate assessment of severity is essential for neurosurgical decision-making. This study aimed to develop and evaluate a fully automated, deep learning-based tool for the standardized assessment of ICH severity, based on the segmentation of the hemorrhage and intracranial structures, and the computation of an objective severity index.

**Methods:** Non-contrast cranial CT scans from patients with spontaneous or traumatic ICH were retrospectively collected from public datasets and a tertiary care center. Deep learning models were trained to segment hemorrhages and intracranial structures. These segmentations were used to compute a severity index reflecting bleeding burden and mass effect through volumetric relationships. Segmentation performance was evaluated on a hold-out test cohort. In a prospective cohort, the severity index was assessed in relation to expert-rated CT severity, clinical outcomes, and the need for urgent neurosurgical intervention.

**Results:** A total of 1,110 non-contrast cranial CT scans were analyzed, 900 from the retrospective cohort and 200 from the prospective evaluation cohort. The binary segmentation model achieved a median Dice score of 0.90 for total hemorrhage. The multilabel model yielded Dice scores ranging from 0.55 to 0.94 across hemorrhage subtypes. The severity index significantly correlated with expert-rated CT severity (p < 0.001), the modified Rankin Scale (p = 0.007), and the Glasgow Outcome Scale–Extended (p = 0.039), and independently predicted the need for urgent surgery (p < 0.001). A threshold ∼300 was identified as a decision point for surgical management (AUC = 0.83).

**Conclusion:** We developed a fully automated and openly accessible pipeline for the analysis of non-contrast cranial CT in intracranial hemorrhage. It computes a novel index that objectively quantifies hemorrhage severity and is significantly associated with clinically relevant outcomes, including the need for urgent neurosurgical intervention.

## INTRODUCTION

Spontaneous and traumatic intracranial hemorrhages (ICH) are life-threatening neurological emergencies associated with high rates of mortality and long-term disability. The global incidence of spontaneous ICH is estimated at 24.6 per 100,000 person-years^1^. In comparison, traumatic brain injury (TBI)–related hemorrhages are even more frequent, with an estimated global incidence of 369 cases per 100,000 population, particularly affecting older adults and polytrauma patients ^2^. Prompt and accurate identification of intracranial bleeding is essential for early clinical decision-making and is routinely performed using non-contrast cranial computed tomography (CT), the imaging modality of choice in the acute setting due to its speed, availability, and sensitivity to blood ^3^.

Over the past decade, artificial intelligence (AI) has driven substantial advances in medical imaging, with numerous deep learning approaches demonstrating expert-level performance in the automatic detection and segmentation of ICH on CT ^4–13^. These tools have the potential to streamline radiological workflows and provide decision support in time-critical environments ^14^. However, most existing AI solutions have focused on detection, segmentation, and classification of hemorrhages, rather than on an objective estimation of hemorrhage severity. Moreover, very few of these tools are publicly accessible ^8,12,15^, and even fewer are deployable in real-world clinical settings without extensive computational infrastructure or technical expertise. This creates a significant translational gap, especially for front-line clinicians such as neurologists, neurosurgeons, emergency physicians, and radiologists.

Yet despite the growing success of automatic hemorrhage segmentation methods, a critical limitation remains in that hemorrhage volume alone does not adequately capture the clinical severity of ICH. The overall impact of a hemorrhage on neurological function is influenced by additional factors, including its subtype (e.g., subarachnoid, intraparenchymal, subdural), anatomical location (e.g., lobar vs. cerebellar), and the degree of associated mass effect ^1^. According to the Monro-Kellie doctrine ^16^, intracranial volume is a fixed space composed of brain parenchyma, cerebrospinal fluid (CSF), and blood. Hemorrhages disrupt this equilibrium, producing compression of compensatory compartments such as ventricles and subarachnoid cisterns. Established radiological indicators, such as midline shift, third ventricle compression, cisternal effacement, and sulcal obliteration, are critical in assessing severity, yet remain subjective and dependent on expert interpretation. These signs are integrated into widely adopted CT-based grading systems used in neurocritical care to guide triage and surgical decisions ^17,18^. However, their interpretation is subject to interrater variability, particularly in borderline findings, which may limit reproducibility and affect clinical consistency ^19^.

Although previous efforts have attempted to integrate the spatial relationship between hemorrhage and surrounding brain structures ^20^, and several studies have explored quantitative methods for estimating midline shift ^21^, to the best of our knowledge, there is currently no fully automated tool capable of combining these elements into a unified severity index. Existing approaches often focus on isolated components, such as hemorrhage localization or volumetry, whereas comprehensive tools that jointly address hemorrhage subtype classification, quantification of key intracranial compartments, and standardized severity grading have received limited attention with regard to systematic development and validation.

In this study, we aim to address this gap through the development of *CEREBLEED*, a novel tool that combines deep learning and volumetric quantification for the automated assessment of intracranial hemorrhage on non-contrast cranial CT. Most importantly, we propose an objective index that integrates hemorrhage volume, structural displacement, and mass effect. This index will be evaluated for its association with clinically relevant outcomes. To promote transparency and reproducibility, the tool is made freely available through a web-based platform.

## METHODS

Ethical approval was obtained from the CEIm Valladolid Ethics Committee under the references PI-24-614-H and PI-25-320-H. The study followed the Strengthening the Reporting of Observational Studies in Epidemiology (STROBE) guidelines. Data are available from the authors upon reasonable request. The developed tool is publicly available through a web platform (https://geibac.uva.es/).

### Study sample

Non-contrast cranial CT scans from patients with spontaneous or traumatic intracranial hemorrhage were retrospectively collected. The training dataset for hemorrhage segmentation models included cases treated at Río Hortega University Hospital (RHUH), Valladolid, Spain, between January 2017 and December 2024, along with additional annotated datasets from BHSD ^22^, CQ500-SEG ^7^, and the INSTANCE Challenge ^11^. A hold-out testing dataset was defined using a random subset of RHUH cases not included in the training set. Additionally, a subset of patients from the RHUH cohort was randomly selected for the development of brain structure segmentation models.

A prospective clinical evaluation cohort was collected between January 2024 and January 2025 and comprised both normal CT scans and pathological scans with intracranial hemorrhage. A detailed flowchart of dataset allocation is shown in Figure 1.

**Figure 1.**
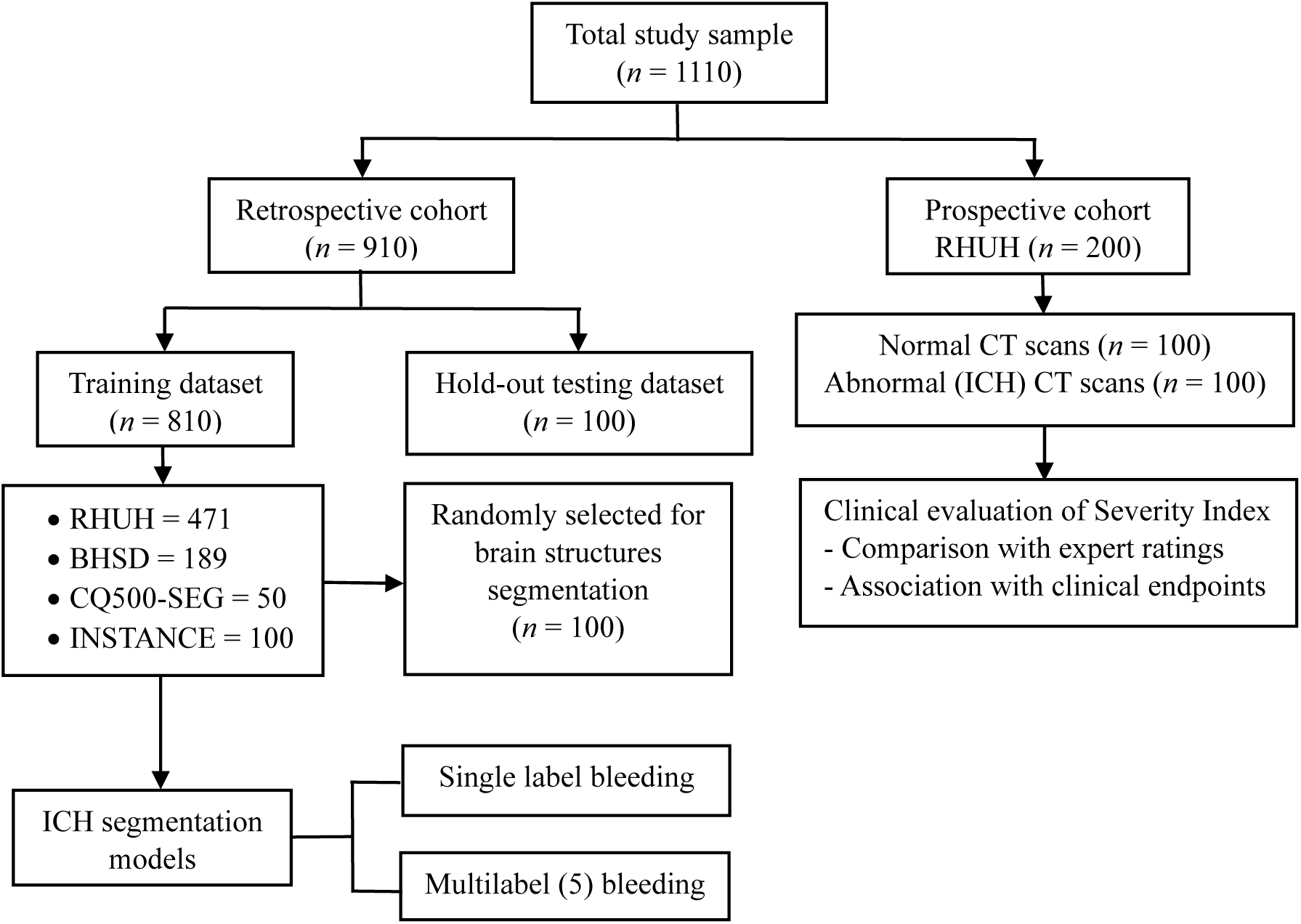
Study design and data flow. ICH= intracranial hemorrhage.

### Image preprocessing

All CT were acquired in Digital Imaging and Communications in Medicine (DICOM) format and converted to Neuroimaging Informatics Technology Initiative (NIfTI) format for preprocessing and model development. Images were registered to a common anatomical reference space to facilitate volumetric quantification and post-processing ^23^.

### Ground truth segmentations for intracranial hemorrhage

Ground truth segmentations of hemorrhage subtypes were manually created for RHUH cases using a semi-automated workflow in ITK-SNAP software (version 4.0.1, http://itksnap.org). Hemorrhages were annotated with unique labels: acute extra-axial hematomas (including epidural and acute subdural)^13^, intraparenchymal hemorrhages, subarachnoid hemorrhages (cisternal and sulcal), intraventricular hemorrhages, and chronic subdural hematomas, encompassing all density patterns ^24^. Segmentations were manually performed by three board-certified neurosurgeons with extensive experience in neuroimaging annotation, as well as in traumatic and neurovascular pathology. All segmentations were subsequently reviewed and approved by a fourth expert neurosurgeon to ensure consistency and accuracy. In the BHSD and CQ500-SEG datasets, the provided multilabel annotations were harmonized with our taxonomy. The INSTANCE dataset, originally containing only binary labels, was manually converted into a multilabel format. All public dataset annotations were reviewed and corrected when necessary, resulting in both multilabel and binary versions of the final dataset.

### Ground truth segmentations for brain structures

A stratified random subsample of the training cohort, balanced across hemorrhage subtypes, was selected to generate ground truth annotations of relevant intracranial anatomical structures. Initial segmentations were generated using the MONAI Auto3DSeg framework ^25^ integrated within 3D Slicer (version 5.7.0) [https://www.slicer.org/], then manually refined. Annotated structures included the midbrain, subarachnoid space, venous sinuses, septum pellucidum, cerebellum, brain parenchyma, ventricular system, and hemorrhage. Anatomical templates derived from an atlas were used to delineate additional key regions, including the basal cisterns, posterior fossa, and third and fourth ventricles. The spatial overlap between hemorrhage masks and these anatomical templates was used to compute region-specific volumetric measures.

### Segmentation model training

Three models were trained using the nnU-Net framework ^26^ employing its default configurations for 2D, 3D low-resolution, and 3D full-resolution modes. One model was trained for binary segmentation of total hemorrhage, another for multilabel classification of hemorrhage subtypes, and a third for segmentation of intracranial anatomical structures. Training employed five-fold cross-validation over 1000 epochs per fold. The loss function combined Dice and cross-entropy. Data augmentation included random rotations, scaling, Gaussian noise and blur, brightness and contrast shifts, gamma corrections, and mirroring. No post-processing was applied to model outputs. Figure 2.

**Figure 2.**
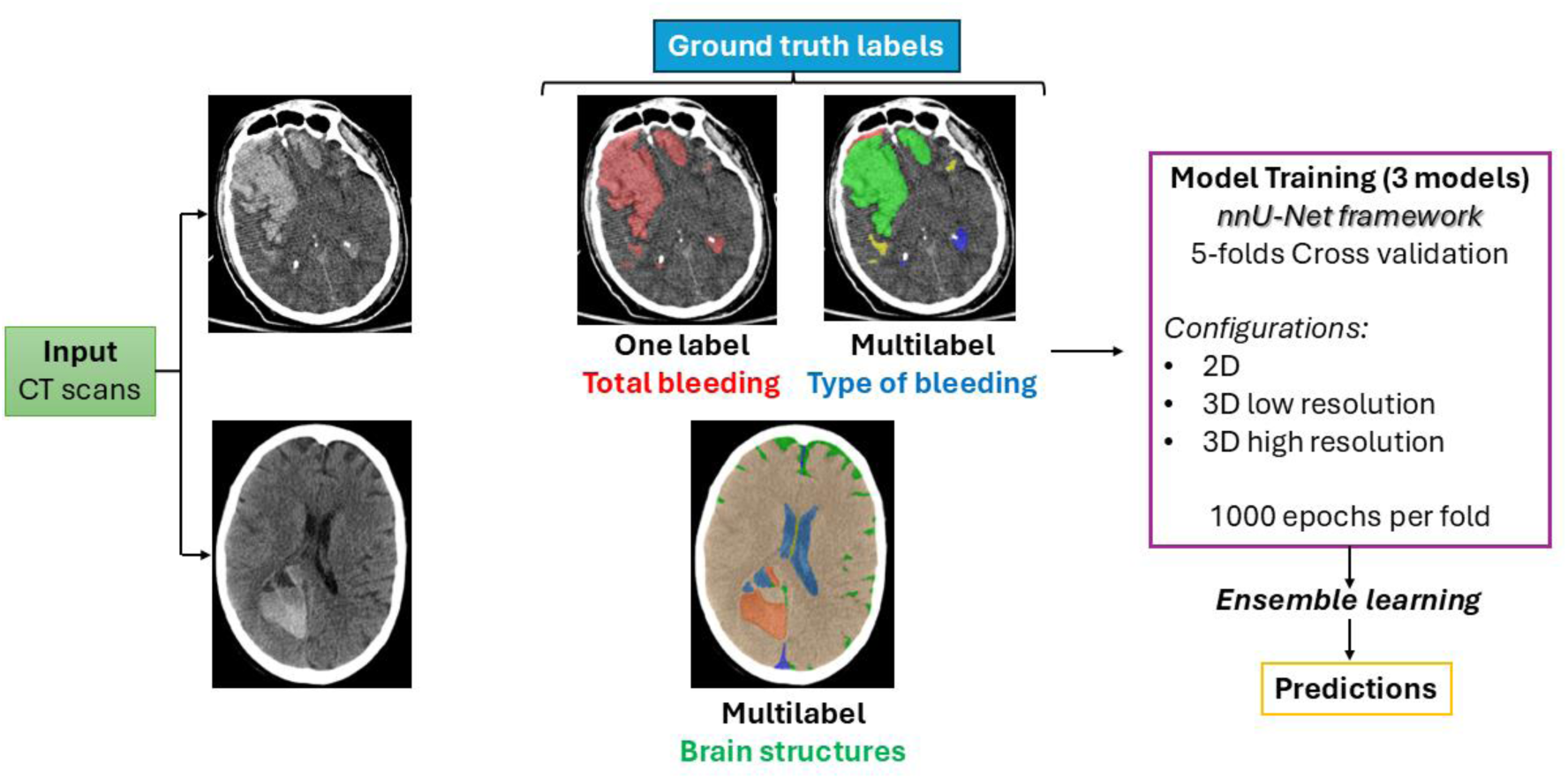
Model training workflow. Input CT scans were annotated with three types of ground truth labels: total bleeding, bleeding subtypes, and brain structures. Three models were trained using the nnU-Net framework with five-fold cross-validation. Final predictions were obtained via ensemble learning.

**Figure 3.**
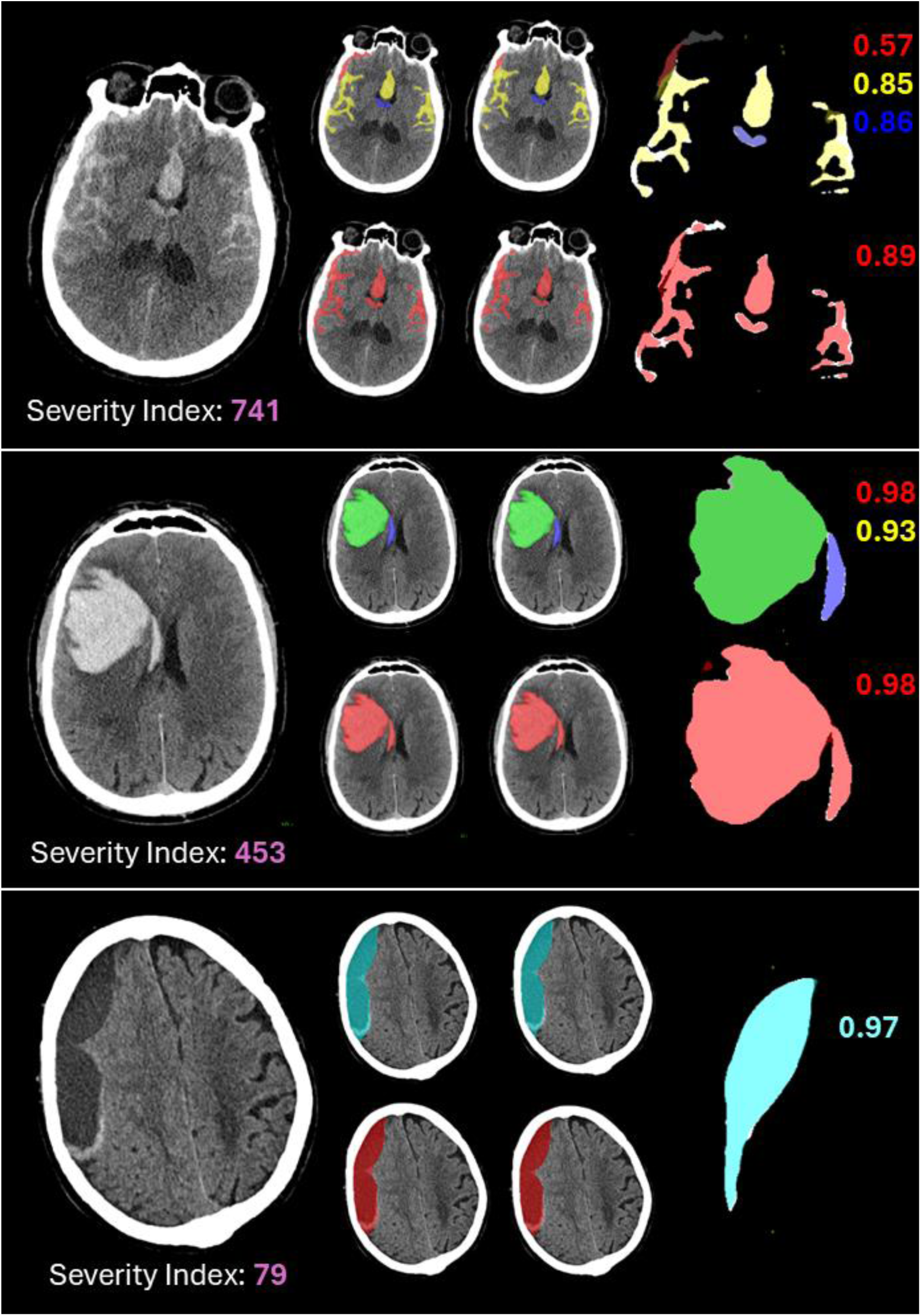
Representative examples of automatic hemorrhage segmentation and severity index calculation. Each row shows a clinical case: (top) aneurysmal subarachnoid hemorrhage, (middle) spontaneous intraparenchymal hemorrhage with ventricular extension, and (bottom) chronic subdural hematoma. From left to right: (1) native axial CT scan; (2) ground truth segmentations (total hemorrhage as single-label and hemorrhage subtypes as multilabel); (3) model predictions; and (4) overlay visualization of both masks, where ground truth is shown as a solid base and predictions are overlaid in color. Dice scores for each class are shown on the right. The Severity Index is shown below each scan.

### Calculation of the hemorrhage severity index

A severity index was computed using mathematical formulas designed to quantify the relationship between hemorrhage volume and the volumes of intracranial structures. These formulas were tailored to each hemorrhage subtype and weighted according to clinical relevance, based on expert consensus from three senior neurosurgeons. For instance, aneurysmal-pattern subarachnoid hemorrhages received higher weights due to their association with acute neurological deterioration and poor outcomes. In contrast, chronic subdural hematomas were assigned lower weights, reflecting their typically indolent clinical course and greater tolerance to large blood volumes resulting from compensatory brain atrophy, which often mitigates mass effect. Additionally, an automated method for quantifying midline shift was developed using the segmentation of midline anatomical landmarks. The specific formulas, weighting coefficients, and midline shift algorithm are protected by intellectual property and are not publicly disclosed.

### Evaluation Metrics

Model performance was evaluated using the USE-Evaluator framework ^27^, which includes volumetric similarity (VS), absolute volume difference (AVD), Dice similarity coefficient (DSC), intersection over union (IoU), Hausdorff distance at the 95th percentile (HD95), average symmetric surface distance (ASSD), and image-level classification metrics such as area under the curve (AUC), correct classification rate (CCR), precision, and specificity.

### Expert Assessment of the Severity Index

In the internal test cohort, the severity index was computed for 100 hemorrhagic cases and independently reviewed by three senior neurosurgeons from the RHUH Neurovascular Unit. Using a consensus-based approach, each case was categorized as “mild,” “moderate,” or “severe” based on perceived mass effect, suspected etiology, and the urgency of surgical intervention.

To assess the correlation between the automated severity index and expert clinical judgment, the index values were compared across the three categories using the Kruskal– Wallis test. A statistically significant difference among groups was observed, supporting the index’s ability to reflect clinically meaningful severity stratification.

### Clinical Utility Evaluation

To assess the clinical utility of the hemorrhage severity index, we conducted a comprehensive evaluation within the prospective validation cohort (n = 200), which included both normal and hemorrhagic CT scans. The index was initially assessed as a binary classifier by comparing its values between patients who did and did not require urgent neurosurgical intervention.

To determine its independent predictive value, a multivariable logistic regression model was applied, adjusting for potential confounders such as age and Glasgow Coma Scale (GCS) on admission.

The association between the severity index and functional outcomes was further evaluated using the modified Rankin Scale (mRS) and the Glasgow Outcome Scale– Extended (GOSE). Index values were compared between favorable and unfavorable outcome groups (mRS ≤ 3 vs. > 3; GOSE ≥ 5 vs. < 5) using the Mann–Whitney U test.

Finally, a receiver operating characteristic (ROC) curve analysis was performed to assess the discriminatory performance of the severity index for predicting the need for neurosurgical intervention. The optimal threshold was determined using Youden’s J statistic, and the area under the curve (AUC), sensitivity, and specificity were reported.

### Computational Resources

Model training and inference were carried out on a Linux Ubuntu 24.04.1 LTS system using Python 3.10 and PyTorch 2.5.1, with CUDA 12.4 and CuDNN 9.5.1 for GPU acceleration. The hardware platform included an AMD Ryzen 9 processor, an NVIDIA RTX 4080 GPU with 16 GB VRAM, and 32 GB of system RAM, providing robust computational performance for 3D medical image analysis.

## RESULTS

A total of 1,110 non-contrast cranial CT scans were included in the study, corresponding to 122,909 axial slices. The training dataset comprised 810 scans: 471 from Río Hortega University Hospital (RHUH), 189 from the BHSD dataset, 100 from the INSTANCE Challenge, and 50 from CQ500-SEG. A hold-out test cohort of 100 RHUH patients, not used for training, was reserved to evaluate the performance of the hemorrhage segmentation model and to assess the correlation between the automated severity index and expert ratings. Finally, a prospective clinical validation cohort was assembled at RHUH, including 200 consecutive patients, 100 with confirmed intracranial hemorrhage and 100 with normal cranial CT scans. Table 1 summarizes the imaging characteristics and bleeding volumes for both the training and hold-out testing cohorts across individual data sources, while Table 2 presents the clinical and demographic features of the prospective evaluation cohort.

**Table 1.**
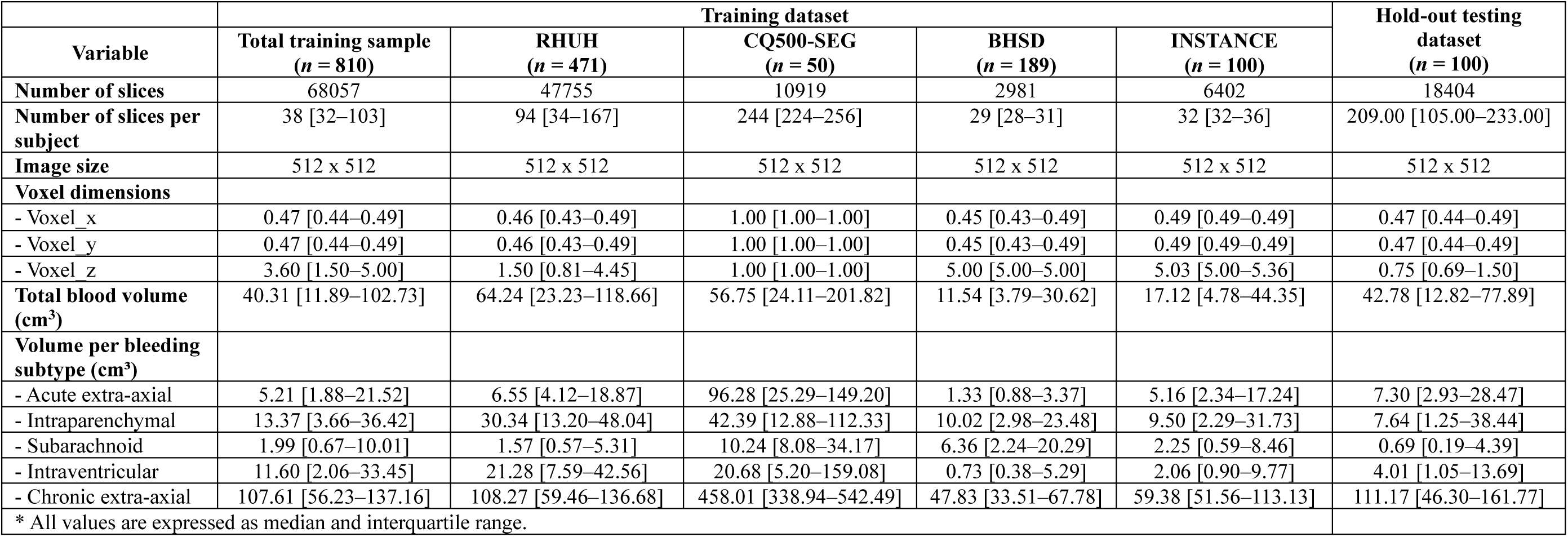
Imaging parameters and bleeding subtype volumes (cm³) across the training dataset and individual data sources.

**Table 2.** Demographics and clinical characteristics of the prospective evaluation cohort.

|  |  |  |
| --- | --- | --- |
| **Variable** | **Abnormal CT (n = 100)** | **Normal CT (n = 100)** |
| Age (years), mean ± SD | 63.44 ± 18.57 | 56.06 ± 20.82 |
| Sex, n (%) |  |  |
| - Male | 54 (54%) | 57 (57%) |
| - Female | 46 (46%) | 42 (42%) |
| GCS at admission, n (%) |  |  |
| - 14–15 | 36 (36%) | 100 (100%) |
| - 9–13 | 35 (35%) | — |
| - $\leq 8$ | 29 (29%) | — |
| mRS at admission, n (%) |  |  |
| - $\leq 3$ | 78 (78%) | 97 (97%) |
| - $> 3$ | 22 (22%) | 3 (3%) |
| Charlson index, n (%) |  |  |
| - 0–2 | 36 (36%) | 58 (58%) |
| - 3–5 | 43 (43%) | 22 (22%) |
| - $> 5$ | 21 (21%) | 19 (19%) |
| Emergency surgery, n (%) |  |  |
| - No | 70 (70%) | — |
| - Yes | 30 (30%) | — |
| mRS at discharge, n (%) |  |  |
| - $\leq 3$ | 47 (47%) | — |
| - $> 3$ | 53 (53%) | — |
| GOSE at discharge, n (%) |  |  |
| - $\geq 5$ | 47 (47%) | — |
| - $< 5$ | 53 (53%) | — |
| Severity index, median (IQR) | 184.6 (84.4–413.5) | — |
| SD = standard deviation; mRS = modified Rankin Scale; GOSE = Glasgow Outcome Scale Extended; IQR = interquartile range. |  |  |

### Segmentation Performance

Segmentation performance metrics for both the binary and multilabel nnU-Net models are summarized in Table 3. The binary model for total hemorrhage segmentation demonstrated excellent overall results in the internal test set, with a median DSC of 0.90 ± 0.01, Jaccard index of 0.82 ± 0.02, and VSI of 0.96 ± 0.01. Precision and recall reached 0.95 ± 0.01 and 0.86 ± 0.02, respectively. Among hemorrhage subtypes, the highest DSC were obtained for intraparenchymal hemorrhage (0.91 ± 0.06) and chronic subdural hematomas (0.94 ± 0.02), while lower values were observed for intraventricular (0.59 ± 0.06) and acute extra-axial hemorrhages (0.55 ± 0.06). Notably, VSI remained consistently high across all subtypes, even in those with modest boundary-level accuracy, highlighting strong volumetric agreement, an essential feature for clinical applicability and severity index computation. The multilabel model for hemorrhage subtype segmentation showed more variable results across classes. DSC were highest for chronic subdural hematoma (0.94 ± 0.02), intraparenchymal hemorrhage (0.91 ± 0.06), and subarachnoid hemorrhage (0.85 ± 0.02). Lower performance was seen in intraventricular hemorrhage (0.59 ± 0.06) and acute extra-axial hematomas (0.55 ± 0.06). Notably, VSI values remained consistently high (>0.93) across all subtypes, including those with lower DSC indicating strong volumetric agreement despite partial discrepancies at the lesion borders.

**Table 3.**
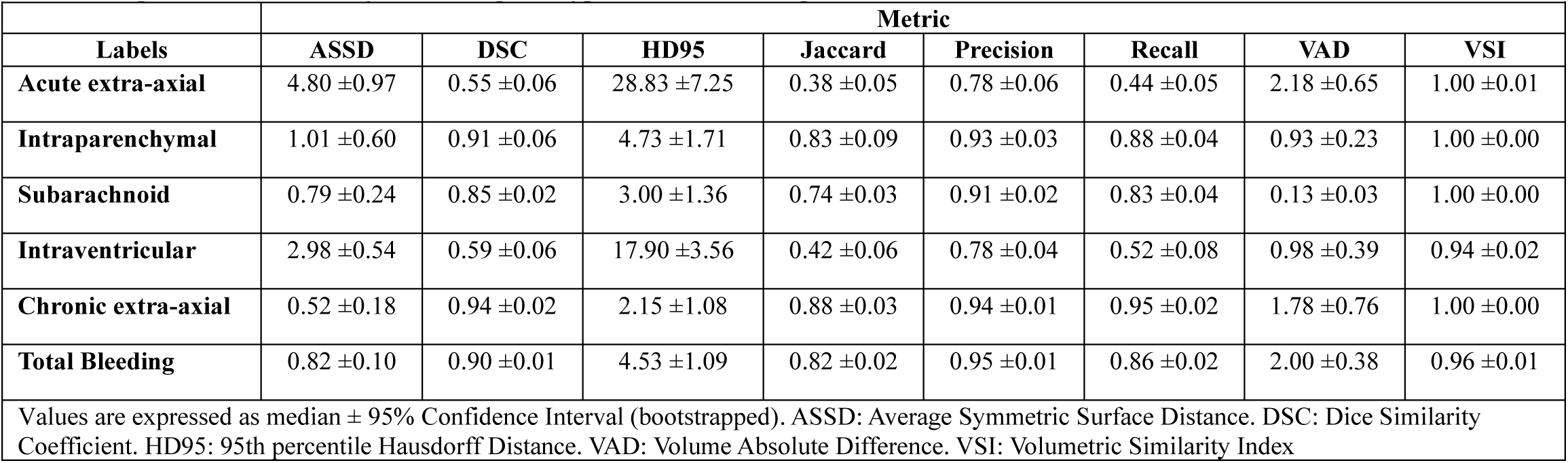
Segmentation metrics by hemorrhage subtype and total bleeding.

|  |  |  |  |  |  |  |  |  |
| --- | --- | --- | --- | --- | --- | --- | --- | --- |
|  | Metric |  |  |  |  |  |  |  |
| Labels | ASSD | DSC | HD95 | Jaccard | Precision | Recall | VAD | VSI |
| Acute extra-axial | 4.80 ±0.97 | 0.55 ±0.06 | 28.83 ±7.25 | 0.38 ±0.05 | 0.78 ±0.06 | 0.44 ±0.05 | 2.18 ±0.65 | 1.00 ±0.01 |
| Intraparenchymal | 1.01 ±0.60 | 0.91 ±0.06 | 4.73 ±1.71 | 0.83 ±0.09 | 0.93 ±0.03 | 0.88 ±0.04 | 0.93 ±0.23 | 1.00 ±0.00 |
| Subarachnoid | 0.79 ±0.24 | 0.85 ±0.02 | 3.00 ±1.36 | 0.74 ±0.03 | 0.91 ±0.02 | 0.83 ±0.04 | 0.13 ±0.03 | 1.00 ±0.00 |
| Intraventricular | 2.98 ±0.54 | 0.59 ±0.06 | 17.90 ±3.56 | 0.42 ±0.06 | 0.78 ±0.04 | 0.52 ±0.08 | 0.98 ±0.39 | 0.94 ±0.02 |
| Chronic extra-axial | 0.52 ±0.18 | 0.94 ±0.02 | 2.15 ±1.08 | 0.88 ±0.03 | 0.94 ±0.01 | 0.95 ±0.02 | 1.78 ±0.76 | 1.00 ±0.00 |
| Total Bleeding | 0.82 ±0.10 | 0.90 ±0.01 | 4.53 ±1.09 | 0.82 ±0.02 | 0.95 ±0.01 | 0.86 ±0.02 | 2.00 ±0.38 | 0.96 ±0.01 |
| Values are expressed as median ± 95% Confidence Interval (bootstrapped). ASSD: Average Symmetric Surface Distance. DSC: Dice Similarity Coefficient. HD95: 95th percentile Hausdorff Distance. VAD: Volume Absolute Difference. VSI: Volumetric Similarity Index |  |  |  |  |  |  |  |  |

Complementary analysis of the binary segmentation model stratified by hemorrhage etiology is detailed in Table 4. The best overall performance was observed for spontaneous intracerebral hemorrhage (IPH), with a DSC of 0.96 ± 0.01, precision of 0.98 ± 0.00, and recall of 0.97 ± 0.01, followed closely by chronic subdural hematoma, which also showed excellent volumetric agreement (VSI: 0.99 ± 0.00). Severe and mild TBI exhibited moderate performance, with DSC values of 0.83 ± 0.04 and 0.88 ± 0.03, respectively. Subarachnoid hemorrhage was the most challenging to segment, with a lower DSC of 0.70 ± 0.08 and recall of 0.59 ± 0.11, likely reflecting its fine, diffuse bleeding patterns.

**Table 4.** Segmentation Performance by Hemorrhage Etiology (Total Bleeding)

|  |  |  |  |  |  |
| --- | --- | --- | --- | --- | --- |
| Metric | sTBI | mTBI | SAH | IPH | cSDH |
|  | 2.05 ±0.54 | 0.82 ±0.24 | 2.48 ±0.86 | 0.35 ±0.13 | 0.27 ±0.08 |
| DSC | 0.83 ±0.04 | 0.88 ±0.03 | 0.70 ±0.08 | 0.96 ±0.01 | 0.97 ±0.01 |
| HD95 | 11.11 ±6.14 | 4.90 ±1.96 | 13.92 ±7.22 | 1.34 ±0.41 | 0.88 ±0.41 |
| Jaccard | 0.71 ±0.06 | 0.79 ±0.04 | 0.54 ±0.09 | 0.92 ±0.02 | 0.94 ±0.02 |
| Precision | 0.88 ±0.02 | 0.95 ±0.01 | 0.87 ±0.03 | 0.98 ±0.00 | 0.97 ±0.01 |
| Recall | 0.80 ±0.07 | 0.83 ±0.04 | 0.59 ±0.11 | 0.97 ±0.01 | 0.97 ±0.01 |
| VAD | 0.93 ±0.05 | 0.92 ±0.02 | 0.79 ±0.18 | 0.99 ±0.01 | 0.99 ±0.00 |
| VSI | 2.05 ±0.54 | 0.82 ±0.24 | 2.48 ±0.86 | 0.35 ±0.13 | 0.27 ±0.08 |
| Values are expressed as median ± 95% Confidence Interval (bootstrapped). ASSD: Average Symmetric Surface Distance. DSC: Dice Similarity Coefficient. HD95: 95th percentile Hausdorff Distance. VAD: Volume Absolute Difference. VSI: Volumetric Similarity Index. sTBI: Severe Traumatic Brain Injury. mTBI: Mild Traumatic Brain Injury. SAH: Subarachnoid Hemorrhage. IPH: Spontaneous Intracerebral Hemorrhage. cSDH: Chronic Subdural Hematoma |  |  |  |  |  |

Image-level classification of hemorrhage subtypes demonstrated consistently high performance across all categories. The highest AUC was achieved for chronic subdural hematoma (0.98 ± 0.01), followed by subarachnoid hemorrhage (0.95 ± 0.02), extra-axial hemorrhage (epidural and acute subdural combined, 0.94 ± 0.03), intraventricular hemorrhage (0.94 ± 0.03), and intraparenchymal hemorrhage (0.93 ± 0.03). Specificity values were also high, all above 0.92, with the highest observed in intraventricular hemorrhage (0.97 ± 0.00). CCR exceeded 89% for all categories, with the best performance in chronic subdural hematoma (90.30% ± 3.00%).

The anatomical structure segmentation model trained on a curated subset of cases yielded a median DSC of 0.81 ± 0.01. Performance by label was as follows: midbrain (0.79 ± 0.01), subarachnoid space (0.76 ± 0.01), venous sinuses (0.80 ± 0.01), septum pellucidum (0.73 ± 0.02), cerebellum (0.85 ± 0.01), brain parenchyma (0.94 ± 0.01), ventricles (0.86 ± 0.01), and hemorrhagic regions (0.75 ± 0.02).

### Correlation Between the Severity Index and Expert Assessment

To evaluate how well the automated severity index aligned with expert clinical judgment, we analyzed 100 cases from the internal RHUH test cohort. Experts categorized each case into three severity levels: mild, moderate, and severe. Median severity index values increased progressively with expert-rated severity: mild (median 30.8, IQR: 20.5–96.2), moderate (101.3, IQR: 82.4–153.9), and severe (244.6, IQR: 151.1–355.6). Differences were statistically significant (Kruskal–Wallis H = 39.6, p < 0.001), confirming that the index accurately reflected clinical severity categories. Figure 4A.

**Figure 4.**
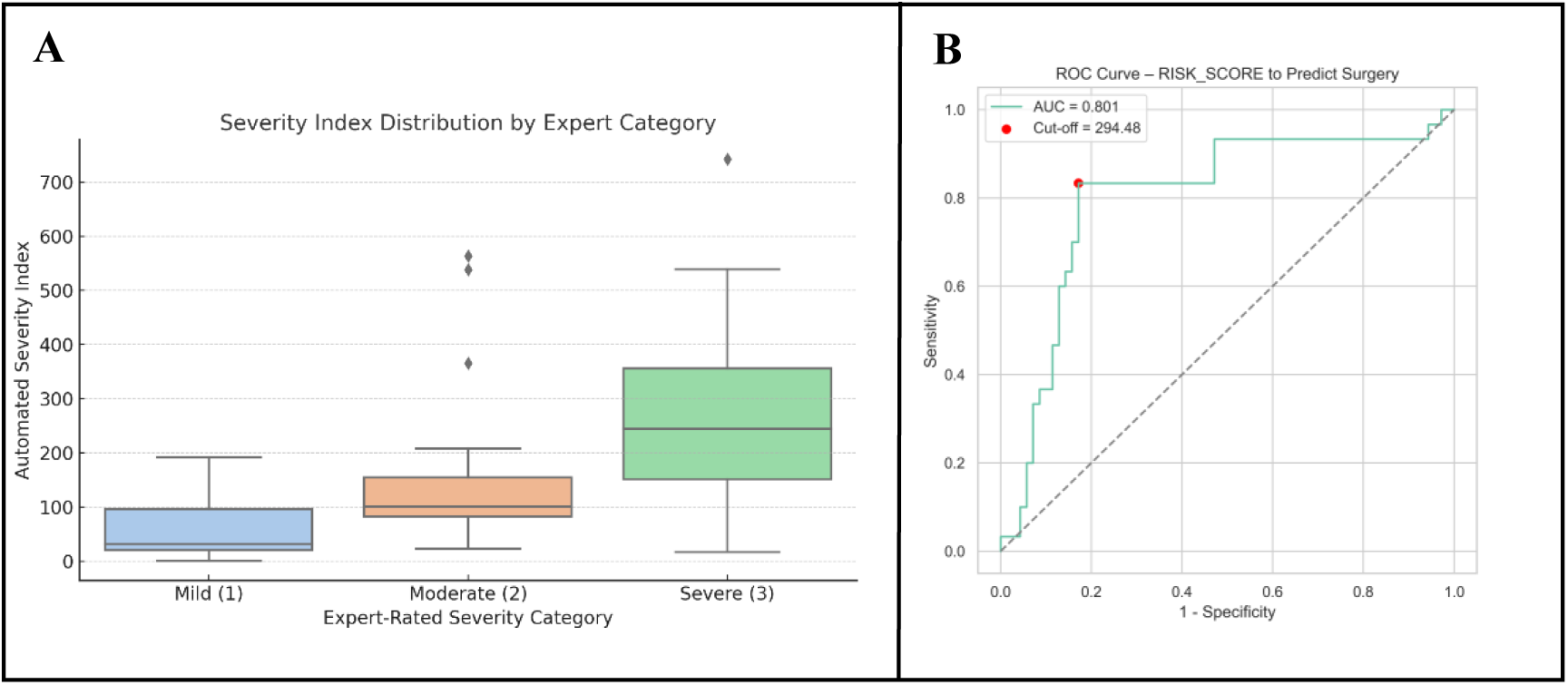
Correlation between the automated severity index and clinical severity endpoints. (A) Distribution of the automated severity index across expert-rated categories (mild, moderate, and severe), demonstrating a clear ascending trend consistent with expert grading. (B) ROC curve for the severity index in predicting the need for emergency surgery, showing an area under the curve (AUC) of 0.80. A threshold of 294.48 yielded an optimal balance between sensitivity and specificity.

### Association Between Severity Index and Clinical Outcomes

Among the 200 patients in the prospective evaluation cohort, all 100 normal CTs received a severity index of 0 and were correctly identified as non-pathological. The hemorrhagic group had a median age of 67 years (IQR: 23.5), with 54% male. On admission, 36% had mild (GCS 14–15), 35% moderate (GCS 9–13), and 29% severe (GCS ≤ 8) neurological impairment. The Charlson Comorbidity Index was 0–2 in 28%, 3–5 in 42%, and >5 in 30%.

The severity index ranged from 5.8 to 1272.4, with a median of 181.6 (IQR: 75.9–241.1). Neurosurgical intervention was required in 30% of cases, while neurological and systemic complications occurred in 39% and 43%, respectively. Functional outcomes varied widely: 23% died (mRS = 6), and 17% recovered fully (mRS = 0); GOSE scores mirrored these findings, with 23% severely disabled or deceased (GOSE = 1) and 19% achieving a score of 8.

Patients with favorable GOSE outcomes (≥5) had lower severity index values (median: 138.1; IQR: 56.1–313.9) than those with poor outcomes (≤4; median: 213.2; IQR: 86.5– 528.2), p = 0.039. A similar association was observed for the modified Rankin Scale: patients with mRS ≤ 3 had lower severity index values (median: 132.6; IQR: 49.3–300.3) than those with mRS ≥ 4 (median: 217.8; IQR: 94.8–528.6), p = 0.007.

Multivariable logistic regression identified the severity index as an independent predictor of neurosurgical intervention (OR: 1.005; 95% CI: 1.002–1.007; p<0.001). Age showed a negative association (OR: 0.97; 95% CI: 0.94–0.99; p = 0.011), indicating lower surgical rates in older patients. GCS was not significantly associated with intervention (p = 0.620).

Receiver operating characteristic (ROC) analysis identified a severity index threshold of 294.5 as optimal for predicting neurosurgical intervention (AUC: 0.83; sensitivity: 0.86; specificity: 0.83), supporting its clinical relevance as a decision-support metric. Figure 4B.

### Automated Clinical Deployment of the Model Pipeline

All trained models have been deployed in an automated pipeline available through the GEIBAC web platform (https://geibac.uva.es/). Users can upload anonymized DICOM images and, within approximately eight minutes, receive segmentation outputs in NIfTI format along with a structured PDF report. This report includes hemorrhage subtype and volume, intracranial structure quantification, and the computed severity index. The system is optimized for real-time clinical settings, offering streamlined decision support for emergency and acute care. Representative examples of the output and clinical interpretation are shown in Figure 5.

**Figure 5.**
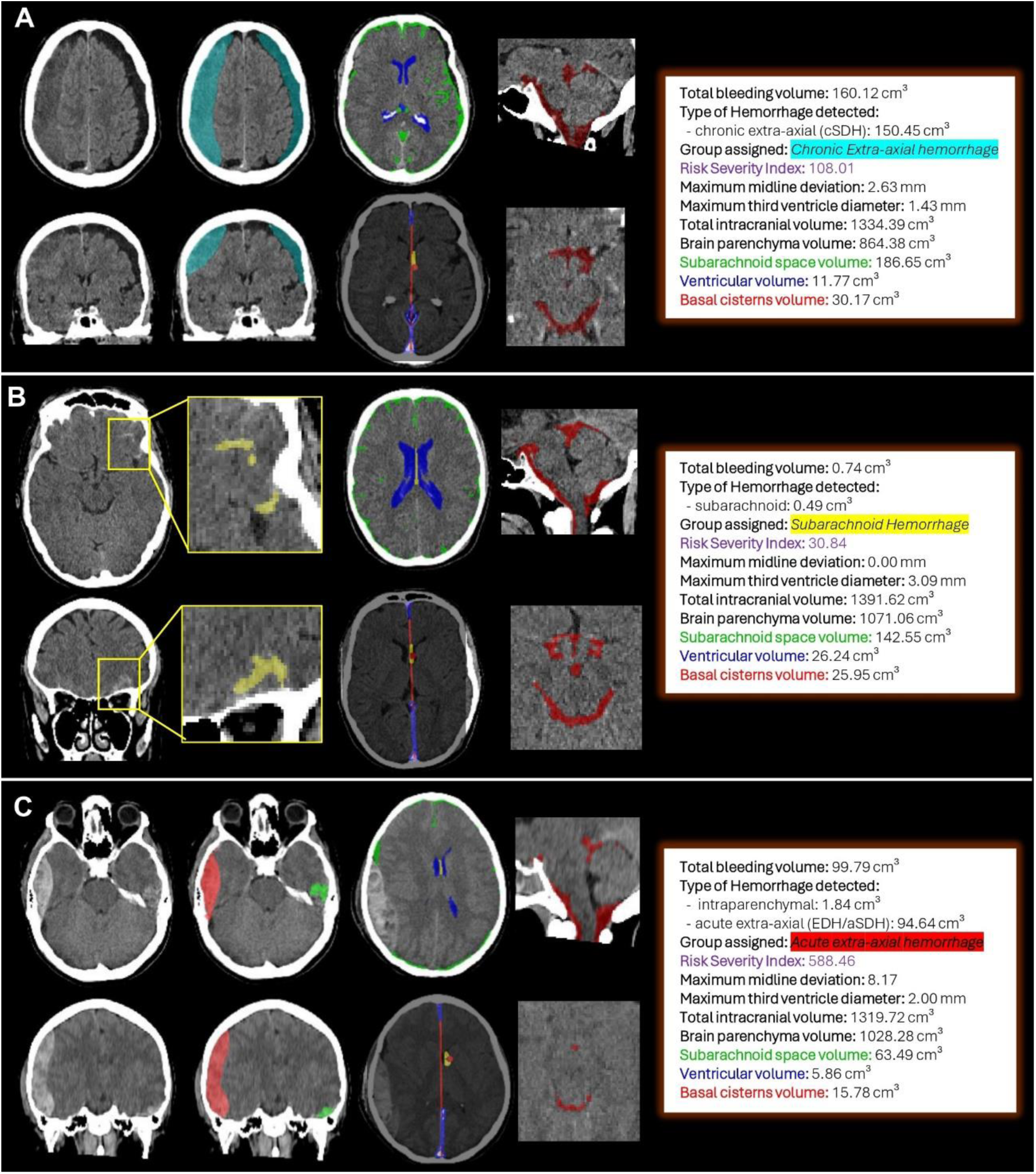
Examples of automated severity index calculation using Cerebleed. From left to right, each panel displays: axial and coronal CT slices showing the hemorrhage with overlaid masks, delineation of intracranial structures and automated midline shift estimation, segmentation of basal cisterns in coronal and axial planes, and the structured report generated by Cerebleed. (A) Chronic subdural hematoma with preserved cisterns and enlarged subarachnoid spaces, likely due to brain atrophy. The calculated severity index was 108.01. The patient did not require emergency surgery but underwent elective evacuation days later. (B) Mild traumatic brain injury with small subarachnoid hemorrhage in the frontal sulci. A low severity index of 30.84 was obtained. The patient was observed for 24 hours and discharged without complications. (C) Severe traumatic brain injury with a large acute right convexity subdural hematoma and left temporal contusion. Diffuse sulcal effacement and basal cistern compression were noted. The severity index reached 588.46. The patient underwent urgent craniotomy.

## DISCUSSION

In this study, we present *CEREBLEED* a fully automated, end-to-end pipeline for the analysis of non-contrast cranial CT scans in patients with intracranial hemorrhage. Our system integrates the full imaging workflow, from preprocessing and global hemorrhage segmentation to subtype classification and volumetric quantification of critical intracranial structures. Most notably, we introduce a standardized severity index derived from volumetric relationships between hemorrhage burden and compensatory anatomical spaces. This index, to our knowledge proposed for the first time, constitutes the principal innovation of our approach, offering an objective and reproducible metric for clinical decision-making in acute care.

In our dataset, both binary and multilabel segmentation models achieved robust performance across a wide spectrum of hemorrhage subtypes. The severity index demonstrated a significant correlation with expert-rated severity and with key clinical outcomes such as mRS and GOSE. Most importantly, it was strongly associated with the need for emergency neurosurgical intervention, underscoring its potential utility in acute clinical decision-making.

While prior studies have demonstrated excellent performance of deep learning models for hemorrhage segmentation ^4,5,12,13^, our approach introduces several unique contributions. Rather than relying on large language models (LLMs) for automated report generation, as proposed in other recent works ^28^, we emphasize a structured, quantitative, and interpretable framework grounded in volumetric analysis. This tool is not intended to replace radiological reports but rather to enhance their objectivity and improve communication among emergency physicians, radiologists, and neurosurgeons.

To ensure robustness and generalizability, we curated a diverse, harmonized training dataset that integrates public and proprietary sources, encompassing both traumatic and spontaneous hemorrhages. All public annotations were reviewed and converted into a consistent multilabel format, allowing standardized learning across data sources.

Importantly, the primary aim of this study is not segmentation per se, which is regarded as an enabling step, but the derivation of a clinically interpretable and structured severity score from the segmentation outputs. Drawing on prior concepts relating hemorrhage volume to cerebrospinal fluid (CSF) space compression ^29^, we developed a metric that captures key indicators of mass effect, including sulcal effacement, cisternal compression, third ventricle narrowing, and midline shift, integrated into a single quantitative index. Unlike conventional visual assessments or the ABC/2 method, which are prone to interobserver variability ^30^, the proposed severity score is objective, reproducible, and integrates both hemorrhage characteristics and the volumetric context of surrounding intracranial structures. It is tailored to hemorrhage subtype and anatomical relationships, providing a more comprehensive and physiologically grounded assessment of severity.

In emergency settings, rapid and accurate interpretation of CT scans is often hampered by variability in expertise and resource availability. Prior studies have documented substantial discrepancies in CT interpretation across radiologists, emergency physicians, and neurosurgeons, which can lead to suboptimal triage and treatment delays, particularly in settings without dedicated neuroradiology support ^31–33^. An objective severity index could support standardized triage, minimize unnecessary interhospital transfers, and facilitate remote consultation in community or rural healthcare settings.

Our tool addresses these challenges by providing a reproducible, quantitative measure that reflects both hemorrhage volume and its effect on intracranial structures and compartments. In our prospective evaluation cohort, the severity index aligned well with expert clinical judgment and independently predicted neurosurgical intervention. A threshold of ∼300 in the severity index was identified as a potential decision cut-off for urgent neurosurgical intervention. While this value showed promising discriminatory capacity in our cohort, further prospective validation is required before clinical adoption.

A key strength of our work is the deployment of the pipeline as a free-access, web-based platform, which eliminates barriers related to hardware, software, or technical expertise. Clinicians can upload anonymized DICOM scans and receive outputs within minutes, including segmentation masks in NIfTI format and a structured PDF report with volumetric analysis and severity index. This implementation supports real-time use, promotes reproducibility, and enhances transparency, addressing major concerns in current biomedical AI research.

Nevertheless, our study has limitations. Although we validated both segmentation and the severity index in internal and prospective clinical cohorts, we did not include an entirely external clinical dataset. However, the integration of multiple public datasets during training contributes to generalizability. We encourage independent external validation and multicenter testing to assess real-world performance. Future research should aim to integrate the severity index into structured radiological reporting systems and assess its impact on clinical workflows and patient outcomes in neurocritical care.

## CONCLUSION

We present a fully automated and freely accessible pipeline for the analysis of non-contrast cranial CT in patients with intracranial hemorrhage. The system performs hemorrhage segmentation, subtype classification, volumetric quantification, and computes a novel index that offers an objective measure of the overall severity of intracranial bleeding. This index demonstrated significant association with clinically relevant outcomes, including the need for urgent neurosurgical intervention. Beyond its predictive value, the tool is designed to support interpretation and enhance communication among physicians involved in the acute management of intracranial hemorrhage.

## Data Availability

All data produced in the present study are available upon reasonable request to the authors.
The developed tool is publicly available through a web platform (https://geibac.uva.es/).

